# A co-produced community engagement workshop in South London to reduce psychosis related stigma

**DOI:** 10.1101/2024.02.21.24303172

**Authors:** Shujun Cai, Joelyn N’Danga Koroma, Charlotte Graham, Meagan McKay, Caroleen Bray, Lauren Colgan, Sarah Lynch, Sharon Fitzell, Isaac Akande, Thomas J Spencer

**Author notes:** Correspondence should be addressed to Dr Thomas Spencer, Department of Psychosis Studies, Institute of Psychiatry, Psychology and Neuroscience, King’s College London, De Crespigny Park, London SE5 8AF, UK.

## Abstract

**Background:** Early access to services is essential for those suffering a psychotic illness or at risk for the condition. Lack of knowledge of services and public stigma are major barriers to care and are associated with increased duration of untreated psychosis and delays in those at clinical high-risk for psychosis (CHR-P) from accessing support. Education and contact programmes have shown promise in reducing stigmatizing views, however, few have been co-produced and delivered by service users and clinical staff. A three-hour co-produced educational workshop was developed and delivered by service users and Outreach and Support in South London (OASIS) staff. The aim was to assess the feasibility of delivering a co-produced workshop to youth community workers, to see if it improved knowledge of psychosis, attitudes to mental health services and reduced stigma towards people with psychosis as well as seeing if the workshop increased community referrals to OASIS.

**Method:** Educational workshops were developed by community stakeholders and service users. Pre- and post-workshop questionnaires assessed knowledge of psychosis, attitudes towards mental health teams and stigma towards individuals with a psychotic disorder. Participants’ and service users’ views on the workshop were explored through two focus groups.

**Results:** 9 workshops were delivered to 75 community participants. Following the workshop participants’ questionnaire scores showed improvements in knowledge of psychosis, improved attitudes towards mental health teams and a reduction in stigmatizing views towards people with psychosis. The workshop was overwhelmingly well received with 97% agreeing with the statement that they learnt something valuable from the workshop. Referrals to the OASIS service increased by 22% following the workshops.

**Conclusion:** The co-produced educational workshop designed and delivered by service users and OASIS staff was successful in improving participants’ knowledge, attitudes and reducing stigma. We observed an increase in the rate of referrals to the OASIS service, although it is difficult to whether this was due to the workshops or to other outreach activities. Future research should examine whether these effects are long-lasting and explore online delivered workshops to reach more participants in the wider community.

## 1. Introduction

Psychotic disorders are debilitating illnesses that can severely impact the individuals, their families and society (Arciniegas, 2015). Research has shown that in many cases patients experience prodromal symptoms before they develop their first episode of psychosis, a period that may last for several years (Klosterkötter et al., 2001). These individuals have a much higher risk of developing psychosis compared to the general population and can be considered as clinical high risk for developing psychosis (CHR-P). Trained practitioners can reliably identify those at CHR-P using standardized semi-structured interviews such as the Comprehensive Assessment of At Risk Mental States (CAARMS; Yung et al., 2005) or the Structured Interview for Psychosis-Risk Syndromes (SIPS; McGlashan et al., 2010). Over the last twenty years there have been major advancements in the detection and prognosis of individuals at clinical high-risk for developing psychosis (Fusar-Poli, et al., 2020) and this approach shows promise for optimizing the benefits of early intervention in psychosis (Fusar-Poli et al., 2017).

Early access to treatment for those experiencing psychosis is essential and is associated with improved outcomes (Correll et al., 2018) whereas delays in detection leads to worse outcomes (Perkins et al., 2005). Furthermore, young people developing psychosis are those least likely to access care through traditional means, and are therefore far more likely to receive treatment only at crisis points (Fusar-Poli, Lai, et al., 2020; Glover & Evison, 2010). Research has shown that lack of knowledge and stigma consititute significant barriers to accessing care through early intervention services (Mueser et al., 2020; Tiller et al., 2023). In their landmark study, Gronholm and colleagues (2017) identified different themes in how stigma may affect pathways to care for CHR-P individuals or those experiencing a first episode of psychosis. From their systematic review, they identified an internalised sense of difference appraised in a negative way, negative reactions from others, a lack of knowledge and feeling labelled or judged by service providers as key aspects of stigma that may be associated with delays in help-seeking and access to care.

Interventions to reduce stigmatizing beliefs about psychosis and the individuals it affects has historically come in many forms: imagined (West et al., 2015) or personal contact with service users (Villani & Kovess - Masfety, 2017), social inclusion programmes (Mazzi et al., 2018), online courses (Cheng et al., 2013), mental health literacy (Sutton et al., 2018), and brief videos (Amsalem, Markowitz, et al., 2021). Overall, meta-analyses confirm small to medium effect sizes for most intervention styles reducing stigma surrounding psychosis (Morgan et al., 2018). Historically, educational interventions have been the most common (Griffiths et al., 2014), with many providing the rationale that stigmatizing beliefs result from a lack of knowledge (Hassab Errasoul et al., 2015; Thornicroft et al., 2008). However, at least for adults, interventions that promote contact with service users, for example a service user sharing their personal recovery story, have been better at reducing stigma (Corrigan et al., 2012).

Educational workshops to reduce stigma around psychosis and improve attitudes to specialist mental health support services have been delivered in the UK (Lloyd-Evans et al., 2015), North America (Blair Irvine et al., 2012; Cheng et al., 2013) and Turkey (Bayar et al., 2009). Results varied between modest and no statistical improvements in both knowledge around psychosis and referral rates to relevant services. None of these interventions were co-produced in partnership with service users and community stakeholders from invention to delivery. Given that effective structural change between communities and services requires service users, community stakeholders and services to work in partnership (Callard et al., 2012; King & Gillard, 2019), this study presents an innovative approach to producing a de-stigmatising public health initiative.

This study aims to assess the feasibility of running co-produced and co-delivered education workshops to increase knowledge of psychosis and reduce stigma about people with psychosis in the London Boroughs of Croydon and Lewisham. The setting for this work was within the Outreach and Support Service in South London (OASIS; Fusar-Poli, Spencer, et al., 2020), a multi-disciplinary community mental health team for young people at CHR-P and part of the pan-London network for psychosis prevention (Fusar-Poli et al., 2019). The target for the workshop intervention was community members working with young people who may have developed a first episode of psychosis or at risk of developing psychosis. The primary aim was to improve participants’ mental health awareness and reduce stigmatizing attitudes towards people with psychosis or those individuals at CHR-P. Secondary aims were to explore community workshop participants’ and service users’ perception of the workshops impact, benefits, limitations, suggested improvements and hopes for educational attainment. We also compared the total number of referrals to the OASIS Croydon and OASIS Lewisham teams before and after the workshop, to see if we saw an increase in referrals from community workers.

## 2. Methods

### 2.1. Workshop Development

The workshops were developed following significant stakeholder engagement and participation. The value of co-production and peer and service user involvement is well documented and found to increase agency, self-esteem and reduce stigma (Mayer & McKenzie, 2017; Sun et al., 2022). The co-production process typically involves principles of reciprocity between partners: valuing the assets of each participant and offering incentivised opportunities for growth, developing participants’ social capital and utilising community networks (Needham & Carr, 2009). With this in mind, clinical staff facilitated a group of eight service users in developing a community education programme based on the Theory of Change project management model (Breuer et al., 2016). One suggestion was to conduct educative workshops to community members. Clinical staff met with prominent community organisations within the Croydon area who echoed a recurrent theme: they recognised their knowledge of psychosis and early warning signs was limited and wanted specialist training to change this. Four OASIS clinicans worked with our community partners, the Croydon BME Forum and Croydon Off The Record, along with six service users to develop central themes of an education workshop. Two further sessions were held where OASIS clinicans and four service users refined the content, structure and activities for the educational workshop.

Once finalized, the workshop was co-delivered with OASIS service users and clinical staff and aimed to address symptoms of psychosis, stigma surrounding psychosis, identification of psychosis in others and awareness of mental health support within the local area. The workshop comprised of multiple teaching methods (see Table 1) to ensure it was engaging and accessible to a variety of learning styles and lasted three hours. All workshops included a powerpoint presentation with a total of 37 slides and was interactive throughout with question and answer sessions, an explanation of the workshop project and time for completion of questionnaires.

**Table 1.** Components of the workshop.

| Teaching aim | Activity | Teaching method |
| --- | --- | --- |
| (a) Symptoms of psychosis | Umbrella of predominant symptoms<br>PHASED acronym<br>(**P**aranoia, **H**allucinations, **A**ged 14-25 years, **S**peech difficulties, **E**xperiencing changes in thoughts, **D**istressed) | Presentation and slide<br>Slide and handout |
| (b) Stigma surrounding psychosis | True / false examples<br>Unusual experiences and prevalence facts | Whole group interaction<br>Breakout group work |
| (c) Identifying psychosis in others | Ice-berg activity<br>Role-play<br>Case studies | Breakout group work and slide<br>Breakout group work and handout<br>Breakout group work, handout and whole group discussion |
| (d) Attitudes towards and accessibility of mental health services | OASIS service user experience<br>Information on psychosis and recovery | Presentation and slide<br>Presentation and slide |

### 2.2. Procedure

The workshop aimed to reach community members working in environments with young people living in the London boroughs of Croydon and Lewisham. Researchers compiled a list of organisations (n=48) deemed to be suitable possible participants. These were identified from previous service development and community engagement activities OASIS had conducted, as well as from stakeholder guidance and directories of local services. An email including brief take-aways from the workshop was sent to managers of identified services and included a flyer with the OASIS logo, website and facts on the prevalence of psychosis. A Continuing Professional Development certificate was included to incentivise attendance. Nine workshops ran over a period of five months in 2019 hosted in a community-based location organised by the OASIS clinical teams. All workshop attendees were given information sheets regarding the nature of the study prior and signed consent forms. No personal identifiable information was recorded and all participant responses were anonymized.

Following the delivery of the workshops, a focus group with community participants, facilitated by OASIS clinical staff who had previously not been involved in the project, was arranged to gather feedback and to evaluate the effectiveness of the workshop. Another focus group was offered to service users who participated in both the development and delivery of the project and was facilitated by staff who knew the service users, upon their request.

### 2.3. Measures

The pre- and post-workshop questionnaires were adapted from Lloyd-Evans et al., (2015), which were originally based on questionnaires by Angermeyer and Matschinger to assess stigma relating to schizophrenia (Angermeyer & Matschinger, 1994, 1996, 1999). Questionnaires were adapted to increase accessibility to the project and shorten the time required for completion (see Supplementary Material for full questionnaires). The questionnaires were divided into four sets of questions to reflect our project aims:

a. Section 1 included 17 items to assess knowledge of behaviours and feelings associated with psychosis. There with four possible responses (Strongly suggests psychosis; Can be part of psychosis; Not usually a feature on psychosis; Not certain) with higher scores indicating better knowledge. Six items containing symptoms not associated with psychosis or unusual beliefs (“red herring” questions) were interspersed with common symptoms. Correct responses for true symptoms (e.g.: feeling persecuted for no clear reason) obtained participants one point if they responded ‘Strongly suggests psychosis’ and ‘Can be part of psychosis’. Correct responses for “red herring” symptoms (e.g.: repetitive checking and cleaning) gained participants a point if they responded ‘Not usually a feature of psychosis’. Possible scores ranged from 0-17 with higher scores indicating better subject knowledge.
b. These questions included 12 items assessing attitudes towards mental health services, rated on a five point Likert Scale with scores ranging from 12-60 (all reverse scored). Higher scores indicated more positive attitudes.
c. These questions included 11 items to assess stigmatized beliefs around people experiencing psychosis or CHR. These statements were on a five point Likert Scale (reverse scored) with possible scores ranging from 11-55. Higher scores indicated more stigmatizing beliefs.
d. These questions assessed identification of psychosis in others. Participants ticked ‘Yes’ or ‘No’ to whether they had worked with someone experiencing psychosis in the past year.

To examine rates of referrals to the OASIS Croydon and OASIS Lewisham teams we compared the number of referrals from the six month period before the workshops July 2018 to December 2018 with the comparable six month period after the workshops, July 2019 to December 2019. Referral sources and whether clients being referred were accepted to the OASIS service were also recorded.

### 2.4. Statistical analysis

Quantitative data were entered into SPSS for Windows version 28 (Chicago, Illinois, USA). Mean scores for the knowledge, stigma and attitudes sub-scales were calculated for both pre- and post-workshop intervention. Difference scores for these paired ordinal data were calculated by subtracting post-intervention means from pre-intervention means. Data were assessed for normality using the Kolmogorov-Smirnoff test of normality. Given that some of the sub-scales were not normally distributed, we assessed the difference scores between pre- and post-workshop questionnaires using Wilcoxon signed rank tests. The focus groups were analysed using realist explicit thematic analysis (Braun & Clarke, 2006). Speech from each focus group was transcribed and coded by one researcher (CB) then refined by other members of the research team, before themes were identified.

## 3. Results

### 3.1 Implementation of the workshop intervention

Nine workshops were delivered and a total of 75 participants attended, their characteristics are reported in Table 2. Participants mainly worked in NHS organizations (32%), third sector organizations (29%), housing services (17%) and school and social care settings (17%) and there were more women (71%) than men.

**Table 2.**
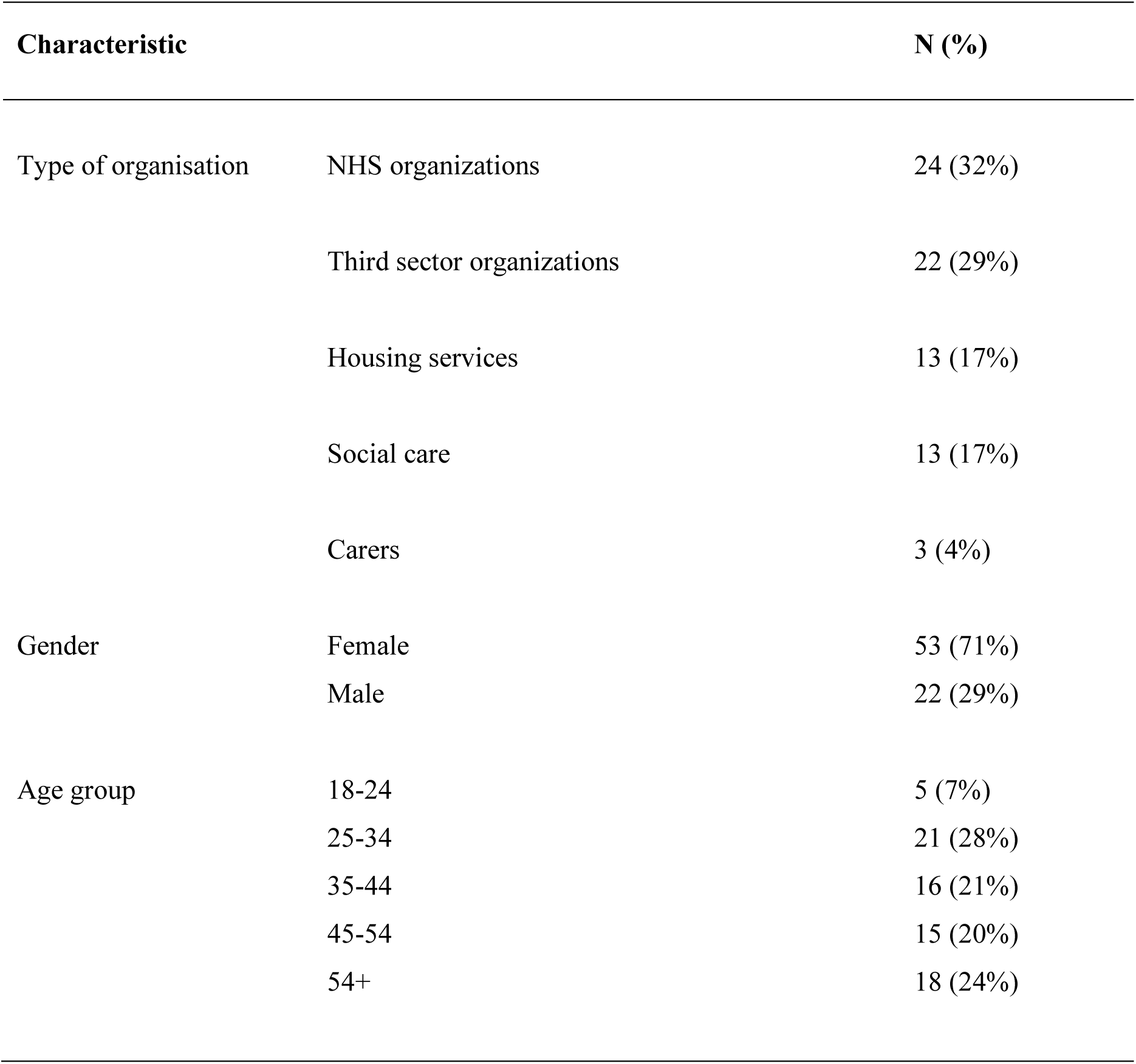
Characteristics of workshop participants.

### 3.2. Evaluation of the workshops

We received complete pre- and post-workshop questionnaires from 74 out of 75 participants (one participant with missing data was removed). The workshop was overwhelmingly well received with 97% agreeing with the statement that they learnt something valuable from the workshop. Wilcoxon matched-pair signed rank tests of pre- and post-workshop scores revealed that participants showed modest increases in their knowledge about psychosis following the workshop (median score increased from 0.64 to 0.82; Z = 6.097, p < 0.001). Similarly, attitudes towards mental health services increased after the workshop intervention (median score increased from 3.83 to 4.08; Z = 4.938, p < 0.001) and stigma reduced (median score reduced from 2.54 to 2.32; Z = -4.888, p < 0.001). Medians for knowledge, attitudes and stigma scores before and after the workshop are are displayed as raincloud plots (Allen et al., 2019) in Figures 1, 2 and 3 respectively.

**Figure 1.**
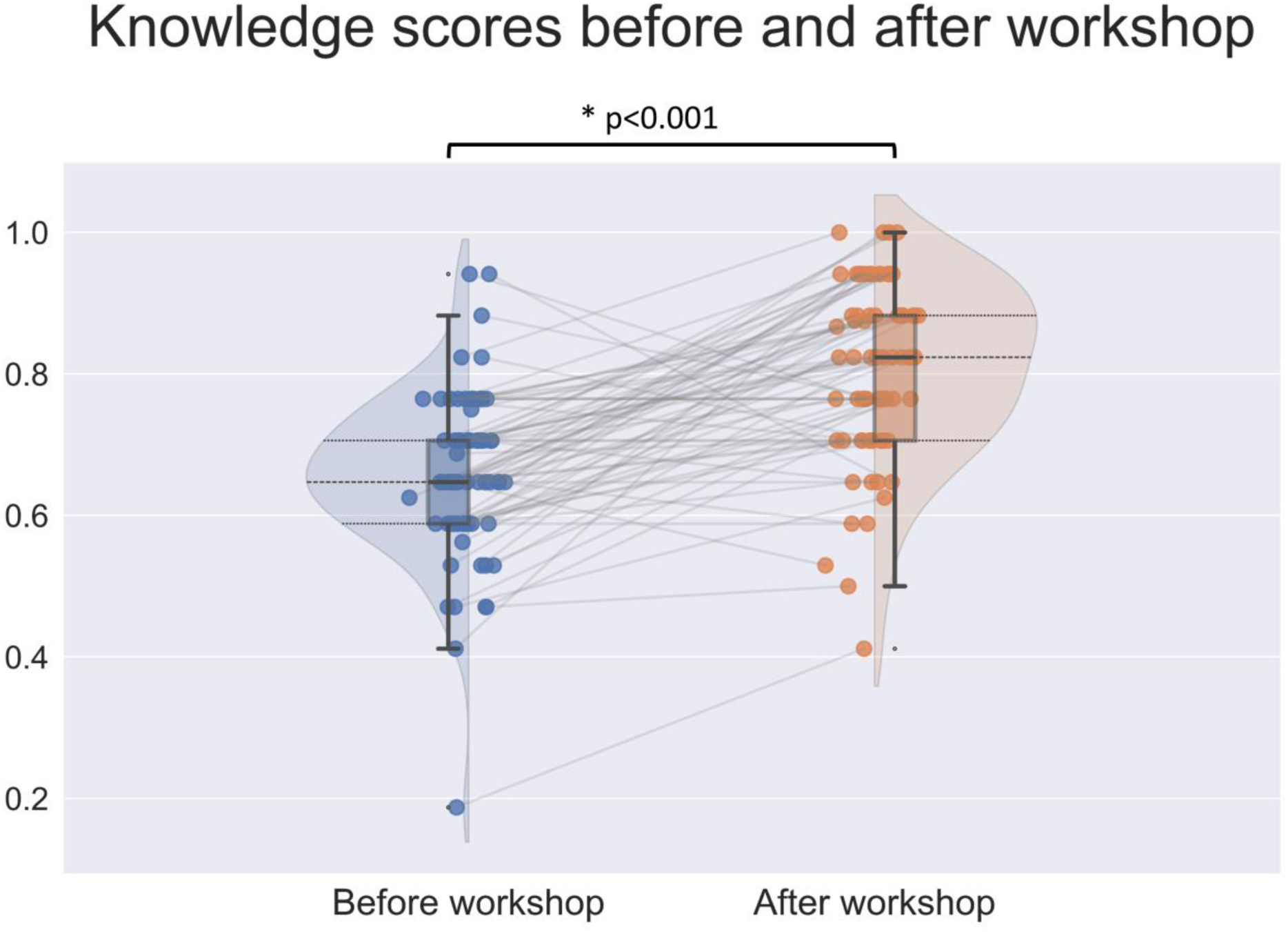
Raincloud plots showing median knowledge questionnaire scores before and after the workshop. Half violin plots show probability density functions and dots show the raw data. Lines between datapoints link the same participants’ before and after questionnaire scores.

**Figure 2.**
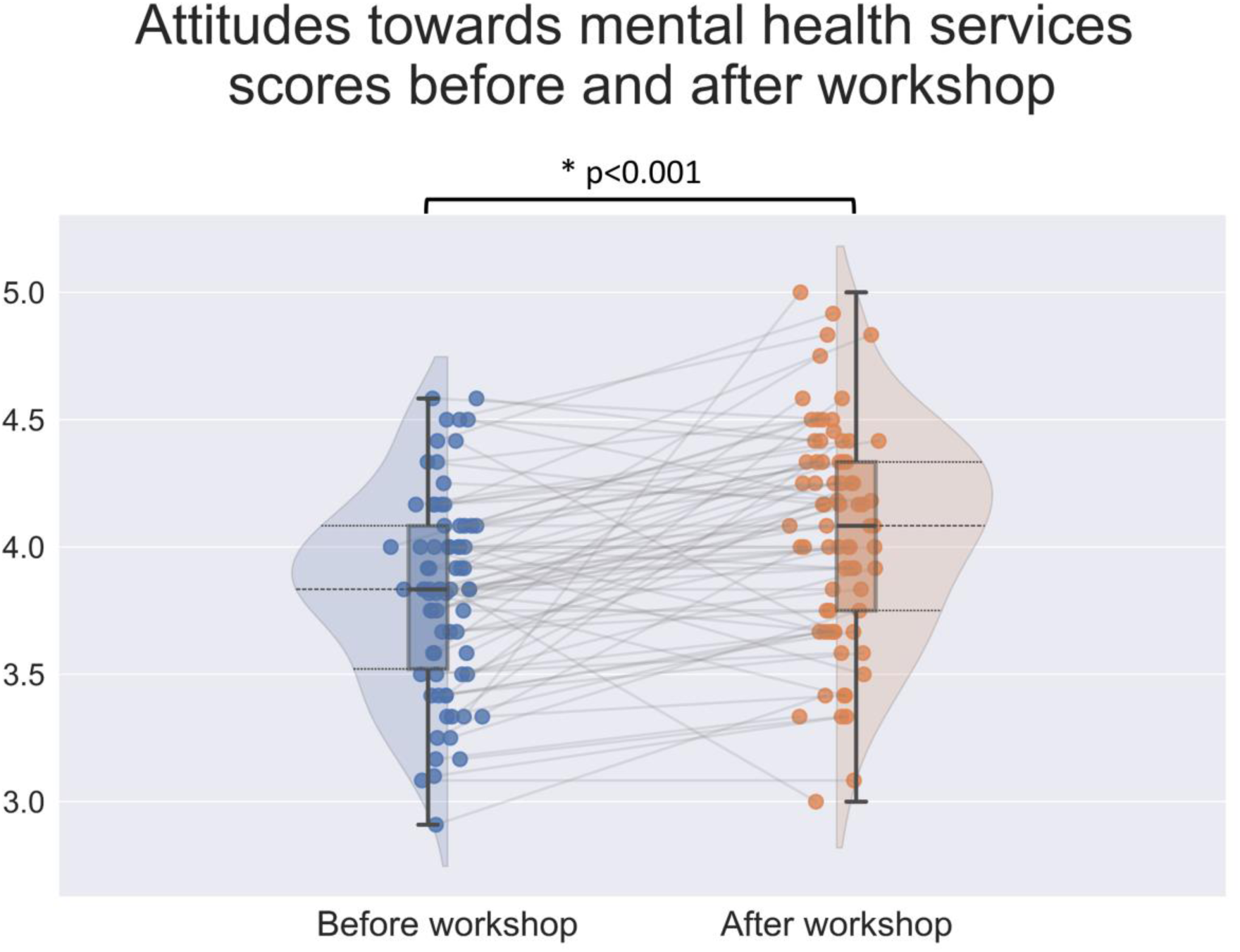
Raincloud plots showing attitudes towards mental health services questionnaire scores before and after the workshop. Half violin plots show probability density functions and dots show the raw data. Lines between datapoints link the same participants’ before and after questionnaire scores.

**Figure 3.**
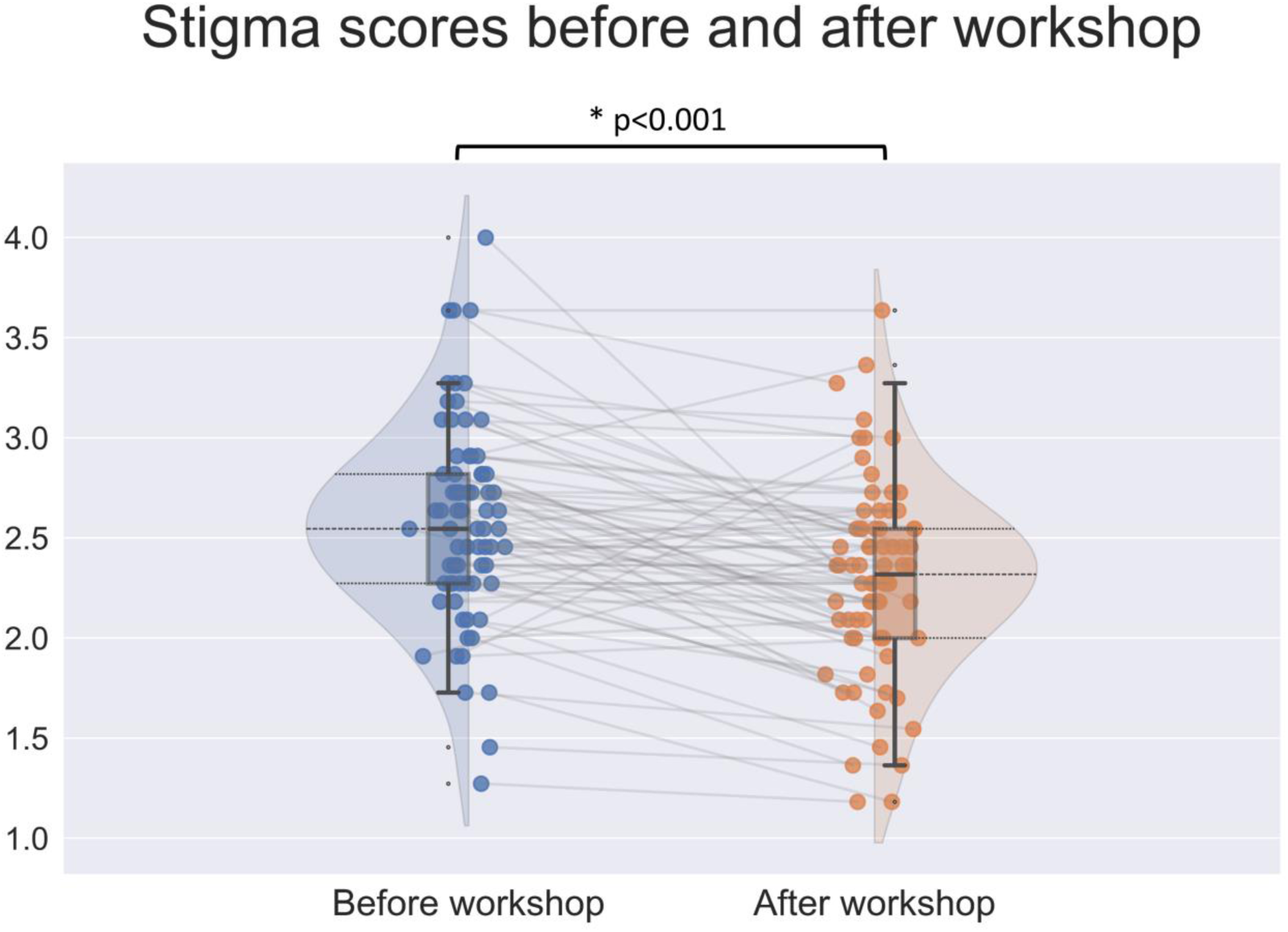
Raincloud plots showing median stigma questionnaire scores before and after the workshop. Half violin plots show probability density functions and dots show the raw data. Lines between datapoints link the same participants’ before and after questionnaire scores.

### 3.3. Participant views of the workshop intervention

The participant focus group was attended by 15 community workers who had taken part in the workshops. Themes were explored around the main research questions: what were the benefits of the workshop, practical limitations and concerns, suggestions for improvements and hopes from educational attainment. All 15 participants endorsed the statement that the workshop was enjoyable and 14 out of 15 participants endorsed the statement that they learnt valuable things from the workshop. Service user collaboration was seen as important, and hearing the service user’s recovery story was rated as the most enjoyable aspect of the workshop experience.

> “All parts of the workshop [were] relevant to the day training. The opening shaped the discussions and the group work. I would suggest that for initial support and supporting people to seek help, this was a good approach”. Workshop participant

Another felt they gained:

> “Understanding of psychosis and early warning signs, as well as what factors can trigger an episode and how common it is. Really enjoyed hearing form a service user”. Workshop participant

Community workers had a concern that people may not want to engage or participate throughout the workshop or that people may not find the benefit of attending. Discussions explored how we could incentivize the community workshops which led to us issuing a continuing professional development certificate to attendees. Pariticpants were keen to learn more about pathways to referral and what happens within the existing pathways of care. Particular clarity around Child and Adolescent Mental Health referrals was sought. Participants appreciated how the workshop began with setting ground rules around confidentiality, which was thought of as a necessary step given the sensitive nature of the topics discussed.

Members of the focus group were keen to have simple and clear definitions and for everyone to begin the workshop by having access to the same framework for language so that the workshop is not filled with jargon. Using clear and simple language will create a more even playing field for everyone to feel their voices are heard. Participants raised the importance of including a Black and Minority Ethinic (BME) focus, especially as people from BME groups face particular challenges in accessing care (Memon et al., 2016; The Sainsbury Centre for Mental Health, 2002), having the Croydon BME forum host workshop sessions helped in this regard. Relatedly, members of the focus group wanted to consider the historical or personal perspectives people may hold regarding mental health including medical, psychological and spiritual. Suggestions included reaffirming that the workshop is being held through a particular lens of understanding. Making this differentiation clear towards the beginning of the workshop may be helpful to state intentions and provide openness to communication.

### 3.4. Service user views of the workshop intervention

The service user focus group was attended by 8 service users. Service users viewed themselves as equally important as staff in the development and delivery of the project, with a focus of all participants being valued and different skill-sets recognized.

> “In some cases we felt more important than staff”. Service user

> “Us adding to conversations was more valued”. Service user

There was a focus on personal outcomes and what matters to the individual. Service users enjoyed talking about their own experiences and being able to impact others with these experiences. Some service users described running the workshops as “cathartic”, helping them to process their emotions surrounding their unusual experiences and getting some distance so that service users could view these critically. Several service users described how the process gave them more confidence.

> “Opened my eyes to my own abilities and what I could do. It developed my confidence and I’m more comfortable with myself”. Service user

A key theme was working in partnership with people service users as catalysts for change. Service users felt “valued” and that the OASIS service “followed through”. There were several suggestions for how to move the project forward including liaising with the Recovery College, or providing workshops to the general public and students at college or university. Service users were happy to be more involved with the planning and running of future workshops and services.

A final theme was the importance of building social capital; helping people become or stay part of their communities. Service users reported that building relationships with other service users has felt “really nice”, “respectful” and “understood”. Another service user reported that working together and doing workshops together had been a “validating” experience and fulfilled a goal “to meet other people who had similar experiences and other like-minded people”. See Table S1 in Supplemental Material for a more detailed breakdown of themes.

### 3.5. Referral rates before and after the workshop

In the period between July 2018 to December 2018, before the workshops were run, OASIS Croydon and OASIS Lewisham received a total of 81 referrals. From July 2019 to December 2019, after the workshops run, OASIS Croydon and OASIS Lewisham received 99 referrals, an increase of 22%.

Examination of whether the referrals were appropriate and accepted onto the OASIS caseload revealed a similar ratio of appropriate referrals for the periods of 2018 and 2019.

## 4. Discussion

### 4.1. Main findings

Following the co-produced workshop, participants’ questionnaire scores showed improvements in knowledge of psychosis, improved attitudes towards mental health teams and a reduction in stigmatizing views towards people with psychosis. These effects were statistically significant, although modest in size. These results replicate findings of similar simple interventions to reduce stigma towards mental health patients in community practitioners in London (Lloyd-Evans et al., 2015), adolescents in a school setting (Dillinger, 2021; Economou et al., 2014; Stuart, 2006) and licensed health care staff (Blair Irvine et al., 2012). The workshop was well-received by the majority of participants. Workshop participants gained great value from hearing the service users’ recovery stories and were able to seek answers to their questions around pathways to care. Service users taking part in the project stressed the importance of working on an equal footing with staff members, gaining confidence and building their social networks.

Referrals to the OASIS Croydon and OASIS Lewisham teams increased by 22% in the six months following the workshops, when the same periods were compared in 2018 and 2019. Referrals were judged to be of a similar quality, as a similar ratio of referrals were accepted onto the OASIS caseload before and after the workshops. It is difficult to argue that this increase in referrals is solely down to the workshops as only a small number of referrals were sent by former workshop participants and it is difficult to control for other service-wide changes and other outreach activities which are a core component of CHR-P teams (Estradé et al., 2022). It was not possible to assess changes in referral rates beyond 2019, as the service was temporarily paused and staff were redeployed during the COVID-19 pandemic. Potentially this kind of educational workshop could help to break down barriers to access mental health care in the community reducing the delay for people developing psychosis to access care. Stigma is a key barrier to accessing mental health care, although a similar intervention was unable to show reductions in duration of untreated psychosis in new referrals to an early intervention service following the community awareness programme (Lloyd-Evans et al., 2015).

### 4.2. Strengths and limitations

As far as we are aware, this is one of the few anti-stigma educational workshops that has been co-produced and co-delivered by service users and clinical staff. Key advantages of this approach are that we were able to embed contact with service users at the heart of the intervention, gaining the benefits of both an educational approach and contact with service users (Corrigan et al., 2012). The study has several limitations, however. We were only able to conduct a relatively small number of workshops (nine) reaching a small cross-section of target population. Larger studies assessing anti-stigma interventions have often utilized online approaches, for example, Blair Irvine and colleagues (2012) were able to recruit 172 licensed health care staff in their online anti-stigma education programme.

Other very promising approaches have adopted videos recorded by service users. Amsalem and colleagues (Amsalem, et al., 2021) created a short online video of a person with schizophrenia describing some of their challenges. This kind of short-form video can easily be shared on social media. By using online recruitment this study was able to assess the impact of watching the video compared with a written form of the content and no intervention. The authors found robust reductions in on stigmatizing views towards people with psychosis, in several stigma domains, due to watching the video compared to the alternative or no intervention (Amsalem, et al., 2021). Furthermore, these effects persisted for 30-days in a subsequent follow-up assessment (Amsalem, Markowitz, et al., 2021). Whist this approach can reach many more participants, it may not be as targeted to address local community-specific barriers to access care as our approach did. By working with local community stakeholders and service users we were able to develop a workshop to reduce stigma and educate community participants on knowledge of psychosis and referral pathways into care. Perhaps the next step is to embrace the online format and create a co-produced online intervention complete with recorded and live components. This would enable us to reach a wider cross-section of the local community in South London and such sessions could more easily be run several times a year to address issues of rapid staff turnover in community organizations.

Another limitation of the study is that we do not have follow-up data to demonstrate whether the effects on knowledge, attitudes towards mental health services and stigma are long-lasting. Several anti-stigma interventions have shown that the effects of the intervention can be long lasting. A one-hour classroom workshop for students on mental illness stigma lead to a reduction in stigma sustained one month after the intervention (Ke et al., 2015). Similarly, a computer-based anti-stigma intervention was able to reduce stigmatizing views and improve mental illness knowledge in graduate students six months after the intervention (Finkelstein et al., 2008). In follow-up studies it would be useful to assess whether the effects of our co-produced workshop on participants’ knowledge, attitudes and stigma persist in the medium or long-term.

### 4.2. Conclusion

A co-produced educational workshop designed and delivered by service users and OASIS staff for community workers in South London lead to improvements in participants’ mental health knowledge and attitudes towards mental health services as well as reducing stigmatizing views towards people with psychosis. The workshops were well received and led to many beneficial outcomes for the service users involved. We observed an increase in the number of referrals to the OASIS Croydon and OASIS Lewisham teams following the workshop, although it is hard to know how much of this was due to the workshop and how much is due to other outreach activities and changes in the service.

## Data Availability

All data produced in the present study are available upon reasonable request to the authors

## Ethics statement

This work was assessed by the South London and Maudsley (SLaM) NHS Foundation Trust Research and Development department, who deemed that the work was a service improvement project and not research, thus it did not need ethical approval.

## Author contributions

CG, SF, IA and TJS designed the study. CG, MM, CB, SL, IA and SF developed and delivered the workshops. SC JNDK CB and TJS analyzed the data. SC and TJS wrote the first draft of the manuscript. CG, SF and TJS acquired funding. All authors contributed to the interpretation of results and to the manuscript.

## Acknowledgements

We thank the community participants, stakeholders and services users who took part and contributed to this study. We also thank the other members of the Outreach and Support in South London (OASIS) team who identified service users and community participants for recruitment into this study.

## Supplementary Materials

**Table S1:**
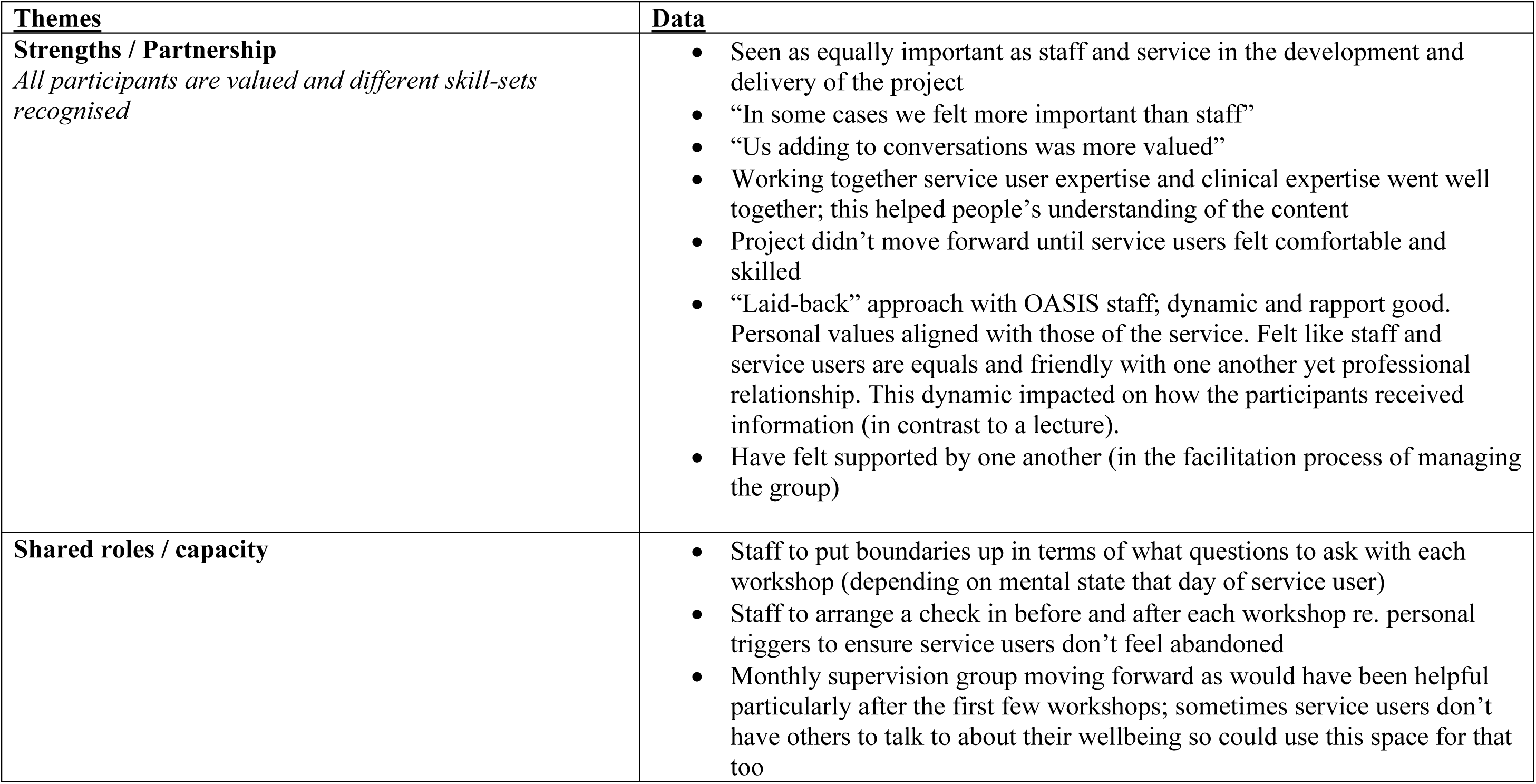
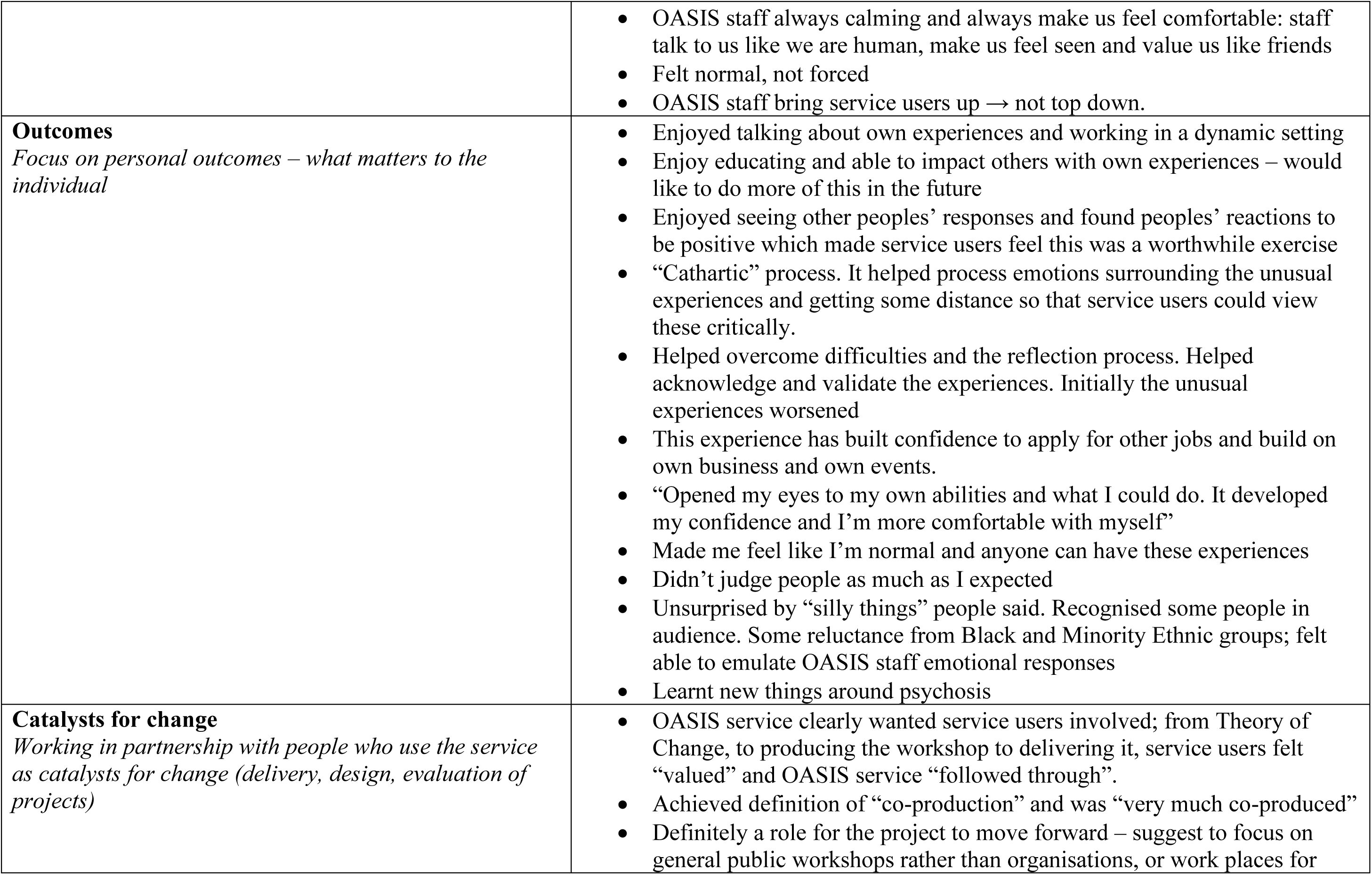
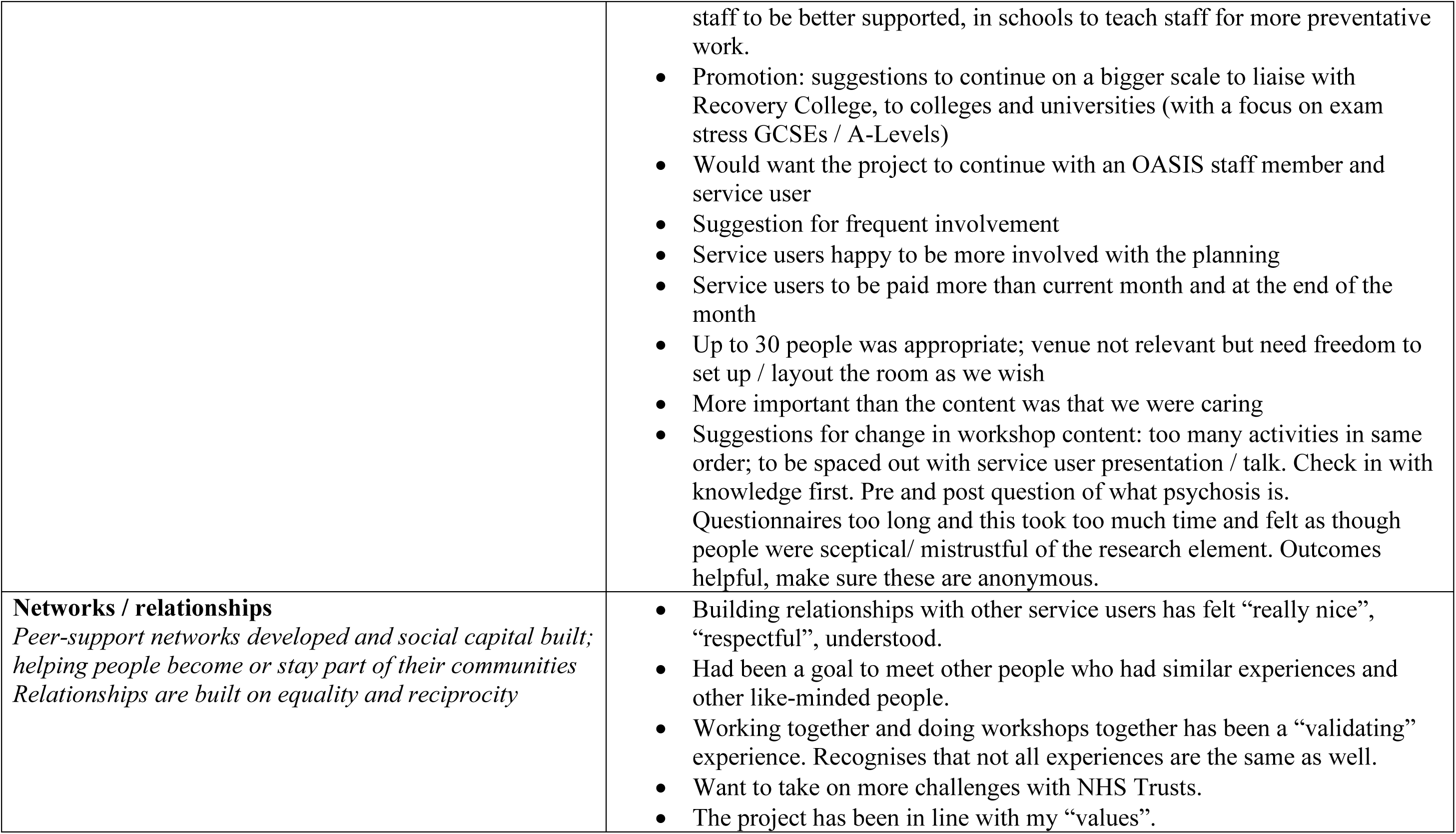
Focus Group Service User Themes.

## References

1. Allen, M., Poggiali, D., Whitaker, K., Marshall, T. R., & Kievit, R. A. (2019). Raincloud plots: a multi-platform tool for robust data visualization. Wellcome Open Research, 4(May), 63. 10.12688/wellcomeopenres.15191.1

2. Amsalem, D., Markowitz, J. C., Jankowski, S. E., Yang, L. H., Valeri, L., Lieff, S. A., Neria, Y., & Dixon, L. B. (2021). Sustained Effect of a Brief Video in Reducing Public Stigma Toward Individuals With Psychosis: A Randomized Controlled Trial of Young Adults. American Journal of Psychiatry, 178(7), 635–642. 10.1176/APPI.AJP.2020.20091293

3. Amsalem, D., Yang, L. H., Jankowski, S., Lieff, S. A., Markowitz, J. C., & Dixon, L. B. (2021). Reducing Stigma Toward Individuals with Schizophrenia Using a Brief Video: A Randomized Controlled Trial of Young Adults. Schizophrenia Bulletin, 47(1), 7–14. 10.1093/schbul/sbaa114

4. Angermeyer, M. C., & Matschinger, H. (1994). Lay beliefs about schizophrenic disorder: the results of a population survey in Germany. Acta Psychiatrica Scandinavica, 89((suppl. 382)), 39–45. 10.1111/j.1600-0447.1994.tb05864.x

5. Angermeyer, M. C., & Matschinger, H. (1996). Public attitude towards psychiatric treatment. Acta Psychiatrica Scandinavica, 94(5), 326–336. 10.1111/j.1600-0447.1996.tb09868.x

6. Angermeyer, M. C., & Matschinger, H. (1999). Lay Beliefs About Mental Disorders. Soc Psychiatry Psychiatr Epidemiol, 34(5), 275–281.

7. Arciniegas, D. B. (2015). Psychosis. CONTINUUM Lifelong Learning in Neurology, 21(3), 715–736. 10.1212/01.CON.0000466662.89908.e7

8. Bayar, M. R., Poyraz, B. Ç., Aksoy-Poyraz, C., & Ankan, M. K. (2009). Reducing mental illness stigma in mental health professionals using a web-based approach. Israel Journal of Psychiatry and Related Sciences, 46(3), 226–230.

9. Blair Irvine, A., Billow, M. B., Eberhage, M. G., Seeley, J. R., McMahon, E., & Bourgeois, M. (2012). Mental illness training for licensed staff in long-term care. Issues in Mental Health Nursing, 33(3), 181–194. 10.3109/01612840.2011.639482

10. Braun, V., & Clarke, V. (2006). Using thematic analysis in psychology. Qualitative Research in Psychology, 3(2), 77–101. 10.1191/1478088706qp063oa

11. Breuer, E., Lee, L., De Silva, M., & Lund, C. (2016). Using theory of change to design and evaluate public health interventions: A systematic review. Implementation Science, 11(1). 10.1186/s13012-016-0422-6

12. Callard, F., Rose, D., & Wykes, T. (2012). Close to the bench as well as at the bedside: Involving service users in all phases of translational research. Health Expectations, 15(4), 389–400. 10.1111/j.1369-7625.2011.00681.x

13. Cheng, C., deRuiter, W. K., Howlett, A., Hanson, M. D., & Dewa, C. S. (2013). Psychosis 101: Evaluating a training programme for northern and remote youth mental health service providers. Early Intervention in Psychiatry, 7(4), 442–450. 10.1111/eip.12044

14. Correll, C. U., Galling, B., Pawar, A., Krivko, A., Bonetto, C., Ruggeri, M., Craig, T. J., Nordentoft, M., Srihari, V. H., Guloksuz, S., Hui, C. L. M., Chen, E. Y. H., Valencia, M., Juarez, F., Robinson, D. G., Schooler, N. R., Brunette, M. F., Mueser, K. T., Rosenheck, R. A., … Kane, J. M. (2018). Comparison of early intervention services vs treatment as usual for early-phase psychosis: A systematic review, meta-analysis, and meta-regression. JAMA Psychiatry, 75(6), 555–565. 10.1001/jamapsychiatry.2018.0623

15. Corrigan, P. W., Morris, S. B., Michaels, P. J., Rafacz, J. D., & Rüsch, N. (2012). Challenging the Public Stigma of Mental Illness: A Meta-Analysis of Outcome Studies. Psychiatric Services, 63(10), 963–973. 10.1176/appi.ps.005292011

16. Dillinger, R. L. (2021). Addressing the Stigma Surrounding Serious Mental Illness in Adolescents: a Brief Intervention. Psychiatric Quarterly, 92(1), 161–167. 10.1007/s11126-020-09787-6

17. Economou, M., Peppou, L. E., Geroulanou, K., Louki, E., Tsaliagkou, I., Kolostoumpis, D., & Stefanis, C. N. (2014). The influence of an anti-stigma intervention on adolescents’ attitudes to schizophrenia: a mixed methodology approach. Child and Adolescent Mental Health, 19(1), 16– 23.

18. Estradé, A., Salazar de Pablo, G., Zanotti, A., Wood, S., Fisher, H. L., & Fusar-Poli, P. (2022). Public health primary prevention implemented by clinical high-risk services for psychosis. Translational Psychiatry, 12(1). 10.1038/s41398-022-01805-4

19. Finkelstein, J., Lapshin, O., & Wasserman, E. (2008). Randomized study of different anti-stigma media. Patient Education and Counseling, 71(2), 204–214. 10.1016/j.pec.2008.01.002

20. Fusar-Poli, P., Estradé, A., Spencer, T. J., Gupta, S., Murguia-Asensio, S., Eranti, S., Wilding, K., Andlauer, O., Buhagiar, J., Smith, M., Fitzell, S., Sear, V., Ademan, A., De Micheli, A., & McGuire, P. (2019). Pan-London Network for Psychosis-Prevention (PNP). Frontiers in Psychiatry, 10(October), 1–10. 10.3389/fpsyt.2019.00707

21. Fusar-Poli, P., Lai, S., Di Forti, M., Iacoponi, E., Thornicroft, G., McGuire, P., & Jauhar, S. (2020). Early Intervention Services for First Episode of Psychosis in South London and the Maudsley (SLaM): 20 Years of Care and Research for Young People. Frontiers in Psychiatry, 11(November), 1–10. 10.3389/fpsyt.2020.577110

22. Fusar-Poli, P., McGorry, P. D., & Kane, J. M. (2017). Improving outcomes of first-episode psychosis: an overview. World Psychiatry, 16(3), 251–265. 10.1002/wps.20446

23. Fusar-Poli, P., Salazar De Pablo, G., Correll, C. U., Meyer-Lindenberg, A., Millan, M. J., Borgwardt, S., Galderisi, S., Bechdolf, A., Pfennig, A., Kessing, L. V., Van Amelsvoort, T., Nieman, D. H., Domschke, K., Krebs, M. O., Koutsouleris, N., McGuire, P., Do, K. Q., & Arango, C. (2020). Prevention of Psychosis: Advances in Detection, Prognosis, and Intervention. JAMA Psychiatry, 77(7), 755–765. 10.1001/jamapsychiatry.2019.4779

24. Fusar-Poli, P., Spencer, T., De Micheli, A., Curzi, V., Nandha, S., & McGuire, P. (2020). Outreach and support in South-London (OASIS) 2001—2020: Twenty years of early detection, prognosis and preventive care for young people at risk of psychosis. European Neuropsychopharmacology, 39, 111–122. 10.1016/j.euroneuro.2020.08.002

25. Glover, G., & Evison, F. (2010). Use of new mental health services by ethnic minorities in England. England: North East Public Health Observatory. Social Care Institute of Excellence.

26. Griffiths, K. M., Carron-Arthur, B., Parsons, A., & Reid, R. (2014). Effectiveness of programs for reducing the stigma associated with mental disorders. A meta-analysis of randomized controlled trials. World Psychiatry, 13(2), 161–175. 10.1002/wps.20129

27. Gronholm, P. C., Thornicroft, G., Laurens, K. R., & Evans-Lacko, S. (2017). Mental health-related stigma and pathways to care for people at risk of psychotic disorders or experiencing first-episode psychosis: A systematic review. Psychological Medicine, 47(11), 1867–1879. 10.1017/S0033291717000344

28. Hassab Errasoul, A., Sutton, M., Doran, C., Robertson, G., Fenlon, N. P., Turner, N., & Clarke, M. (2015). Creating a curriculum on psychosis: A pilot training programme with youth workers. Early Intervention in Psychiatry, 9(5), 412–421. 10.1111/eip.12158

29. Ke, S., Lai, J., Sun, T., Yang, M. M. H., Wang, J. C. C., & Austin, J. (2015). Healthy Young Minds: The Effects of a 1-hour Classroom Workshop on Mental Illness Stigma in High School Students. Community Mental Health Journal, 51(3), 329–337. 10.1007/s10597-014-9763-2

30. King, C., & Gillard, S. (2019). Bringing together coproduction and community participatory research approaches: Using first person reflective narrative to explore coproduction and community involvement in mental health research. Health Expectations, 22(4), 701–708. 10.1111/hex.12908

31. Klosterkotter, J., Hellmich, M., Steinmeyer, E. M., & Schultze-Lutter, F. (2001). Diagnosing Schizophrenia in the Initial Prodromal Phase. Archives of General Psychiatry, 58, 158–164.

32. Lloyd-Evans, B., Sweeney, A., Hinton, M., Morant, N., Pilling, S., Leibowitz, J., Killaspy, H., Tanskanen, S., Totman, J., Armstrong, J., & Johnson, S. (2015). Evaluation of a community awareness programme to reduce delays in referrals to early intervention services and enhance early detection of psychosis. BMC Psychiatry, 15(1), 1–13. 10.1186/s12888-015-0485-y

33. Mayer, C., & McKenzie, K. (2017). Health Social Care Comm - 2017 - Mayer - it shows that there s no limits the psychological impact of co-production for.pdf. Health and Social Care in the Community, 25(3), 1181–1189. doi: 10.1111/hsc.12418

34. Mazzi, F., Baccari, F., Mungal, F., Ciambellini, M., Brescancin, L., & Starace, F. (2018). Effectiveness of a social inclusion program in people with non-affective psychosis. Community Mental Health Journal, 18, 179. 10.1007/s10597-020-00547-1

35. McGlashan, T., Walsh, B., & Woods, S. (2010). The psychosis-risk syndrome: Handbook for diagnosis and follow-up. Oxford University Press.

36. Memon, A., Taylor, K., Mohebati, L. M., Sundin, J., Cooper, M., Scanlon, T., & De Visser, R. (2016). Perceived barriers to accessing mental health services among black and minority ethnic (BME) communities: A qualitative study in Southeast England. BMJ Open, 6(11), 1–9. 10.1136/bmjopen-2016-012337

37. Morgan, A. J., Reavley, N. J., Ross, A., Too, L. S., & Jorm, A. F. (2018). Interventions to reduce stigma towards people with severe mental illness: Systematic review and meta-analysis. Journal of Psychiatric Research, 103(March), 120–133. 10.1016/j.jpsychires.2018.05.017

38. Mueser, K., DeTore, N., Kredlow, M., Bourgeois, M., Penn, D., & Hintz, K. (2020). Clinical and demographic correlates ofstigma in first-episode psychosis: the impactof duration of untreated psychosis. Acta Psychiatrica Scandinavica, 141, 157–166.

39. Needham, C., & Carr, S. (2009). Emerging evidence base for adult social care transformation. Social Care Institute of Excellence, March. http://www.scie.org.uk/publications/briefings/files/briefing31.pdf

40. Perkins, D. O., Hongbin, G., Boteva, K., & Lieberman, J. A. (2005). Relationship Between Duration of Untreated Psychosis and Outcome in First-Episode Schizophrenia : A Critical Review and Meta-Analysis. American Journal of Psychiatry, 162, 1785–1804.

41. Stuart, H. (2006). Reaching out to high school youth: The effectiveness of a video-based antistigma program. Canadian Journal of Psychiatry, 51(10), 647–653. 10.1177/070674370605101004

42. Sun, J., Yin, X., Li, C., Liu, W., & Sun, H. (2022). Stigma and Peer-Led Interventions: A Systematic Review and Meta-Analysis. Frontiers in Psychiatry, 13(July), 1–12. 10.3389/fpsyt.2022.915617

43. Sutton, M., O’Keeffe, D., Frawley, T., Madigan, K., Fanning, F., Lawlor, E., Roche, E., Kelly, A., Turner, N., Horenstein, A., O’Callaghan, E., & Clarke, M. (2018). Feasibility of a psychosis information intervention to improve mental health literacy for professional groups in contact with young people. Early Intervention in Psychiatry, 12(2), 234–239. 10.1111/eip.12410

44. The Sainsbury Centre for Mental Health. (2002). Breaking the Circles of Fear. A review of the relationship between mental health services and African and Caribbean communities. (Vol. 33, Issue 4).

45. Thornicroft, G., Brohan, E., Kassam, A., & Lewis-Holmes, E. (2008). Reducing stigma and discrimination: Candidate interventions. International Journal of Mental Health Systems, 2, 1–7. 10.1186/1752-4458-2-3

46. Tiller, J., Maguire, T., & Newman-Taylor, K. (2023). Early intervention in psychosis services: A systematic review and narrative synthesis of barriers and facilitators to seeking access. European Psychiatry, 66(1). 10.1192/j.eurpsy.2023.2465

47. Villani, M., & Kovess - Masfety, V. (2017). Could a short training intervention modify opinions about mental illness? A case study on French health professionals. BMC Psychiatry, 17(1), 1–9. 10.1186/s12888-017-1296-0

48. West, K., Turner, R., & Levita, L. (2015). Applying imagined contact to improve physiological responses in anticipation of intergroup interactions and the perceived quality of these interactions. Journal of Applied Social Psychology, 45(8), 425–436. 10.1111/jasp.12309

49. Yung, A. R., Yuen, H. P., McGorry, P. D., Phillips, L. J., Kelly, D., Dell’Olio, M., Francey, S. M., Cosgrave, E. M., Killackey, E., Stanford, C., Godfrey, K., & Buckby, J. (2005). Mapping the onset of psychosis: The Comprehensive Assessment of At-Risk Mental States. Australian and New Zealand Journal of Psychiatry, 39(11–12), 964–971. 10.1111/j.1440-1614.2005.01714.x

